# Emergence of the novel SARS-CoV-2 lineage P.4.1 and massive spread of P.2 in South Brazil

**DOI:** 10.1101/2021.04.14.21255429

**Authors:** Fernando Hayashi Sant’Anna, Ana Paula Muterle Varela, Janira Prichula, Juliana Comerlato, Carolina Baldisserotto Comerlato, Vinicius Serafini Roglio, Gerson Fernando Mendes Pereira, Flávia Moreno, Adriana Seixas, Eliana Márcia Wendland

## Abstract

South Brazil has been the novel epicenter of Coronavirus Disease 2019 (COVID-19) in 2021, accounting for the greatest number of cumulative cases and deaths (per 100 thousand inhabitants in a week) worldwide. In this study, we analyzed 340 whole genomes of SARS-CoV-2, which were sampled between April and November 2020 in 33 cities in South Brazil. We demonstrated the circulation of two novel emergent lineages, described here as P.4 and P.4.1 (provisionally termed VUI-NP13L), and seven lineages that had already been assigned (B.1.1.33, B.1.1.28, P.2, B.1.91, B.1.1.94, B.1.195 and B.1.212). P.2 and P.4.1 demonstrated massive spread from approximately September/October 2020. Constant and consistent genomic surveillance is crucial to identify newly emerging SARS-CoV-2 lineages in Brazil and to guide decision making in the Brazilian Public Healthcare System.

## Introduction

Since the emergence of SARS-CoV-2 (severe acute respiratory syndrome coronavirus 2) in China at the end of 2019, COVID-19 (coronavirus disease 2019) has been responsible for more than 2.7 million deaths worldwide^1–3^.. Brazil is second in terms of the number of COVID-19 deaths, only behind the USA, registering more than 303,462 deaths and 12,320,169 cumulative cases as of 26 March 2021. Rio Grande do Sul (RS), the most southern state of Brazil, borders Argentina and Uruguay and had its first COVID-19 case confirmed at the end of February 2020. More than one year later, RS reached a peak of infections leading to a collapse of the health system^4,5^, with 14,957 deaths and 742,866 cases^6,7^.

Even after one pandemic year and the introduction of SARS-CoV-2 vaccination worldwide, we have no forecasted end of the COVID-19 pandemic. Additionally, resurgence of COVID-19 after the first wave and after reaching high seroprevalence may drive positive selection of new lineages^8,9^. Constant epidemiological genomic surveillance through large-scale pathogen genome sequencing has played a major role in the detection and spatial-temporal distribution of SARS-CoV-2 lineages. Therefore, these data allow quasi-real-time tracking of viral dynamics around the globe^10–15^.

Despite the volume of COVID-19 cases in Brazil, a task force created by the scientific community was able to sequence only approximately 0.03% of all positive SARS-CoV-2 cases through the pandemic’s first year^16,17^. To date, 59 different lineages were announced in Brazil, being the majority sequenced in São Paulo, Rio de Janeiro, Rio Grande do Sul, and Amazonas^16^. The lineages more frequently identified and currently in circulation are B.1, the first ancestral lineage introduced in Brazil, B.1.1.212, B.1.1.33, B.1.1.74, B.1.1.28, B.1.1.143, B.1.1.94, and three recently assigned lineages, P.1, P.2, and N.9, derived from B.1.1.28 and B.1.1.33^16,18^. From November 2020 until now, P.1 and P.2 have been widely represented and distributed across all Brazilian regions. Recently, in Rio Grande do Sul, phylogenetic analysis revealed the occurrence of a novel cluster from B.1.1.28, provisionally named VUI-NP13L^17^.

Despite sequencing efforts, there is no accurate knowledge about the frequency of SARS-CoV-2 lineage distribution in Brazil, mainly due to sampling gaps in spatiotemporal strata and the low number of genomes^19,20^. At the time of this manuscript (26 March 2021), approximately 4,500 SARS-CoV-2 genomes from Brazil were available in GISAID. This study aimed to reconstruct the spatiotemporal pattern of SARS-CoV-2 spread in the first year of the pandemic in South Brazil, searching for the emergence of novel lineages. Whole-genome sequencing was performed for 340 SARS-CoV-2 genomes, the largest temporal (April and November 2020) monitoring of Brazil to date.

## RESULTS

### Sampling, data acquisition and genome assembly

Between 24 April and 30 November, 2020, 13,700 nasopharyngeal swab samples from four regions of Rio Grande do Sul were laboratory-confirmed to have SARS-CoV-2 infection at Epiclin Laboratory. From those, 353 samples were selected by region and time for whole-genome sequencing. We verified that only seven of 96 consensus sequences of the first sequencing run presented a complete region spanning the position 19204 to 19616, corresponding to amplicon 64 of the ARTIC framework (Supplementary Figure 1). For subsequent sequencing runs, the depth of the amplicon 64 region was increased by lowering the annealing temperature of the multiplex tiling-PCR by 1 °C.

After sequencing, 340 genomes displayed the minimum quality requirements for further analyses. The number of paired-end reads generated per sample varied from 132,396 to 3,480,771. Genome sequences comprised at least 97.44% of the Wuhan-Hu-1 reference genome (GenBank accession number: NC_045512.2), independent of the threshold cycle (Ct), with depth coverage ranging from 554 x to 14,935 x (Supplementary Data 1). These results are associated with the optimization of the sequencing library preparation protocol (Supplementary Fig. 1).

Our sequencing effort was composed of a homogeneous distribution of samples along a temporal window from April to November 2020, when compared with a dataset available for the same period (Supplementary Fig. 2). Our findings more than double the number of SARS-CoV-2 sequences (from 232 to 572) from South Brazil available in GISAID.

### Monitoring of SARS-CoV-2 lineages in South Brazil

Genomes reported in this study were assigned to 12 lineages based on the proposed dynamic nomenclature of Phylogenetic Assignment of Named Global Outbreak Lineages (PANGOLIN)^21,22^ (Supplementary Data 1). To determine the phylogenomic and epidemiologic characteristics of the SARS-CoV-2 genomes, we performed a phylogenomic analysis containing the genomes described in our study combined with a dataset of 3,965 global representative genomes, enriched for South American sequences (1,340 Brazilian genomes; Supplementary Data 2 and 3). Therefore, seven main lineages were distributed in three main clusters: A, B.1.1.28 (n = 116; 34.1%), P.2 (n = 14; 4.1%), B.1.1.94 (n = 1; 0.3%); B, B.1.1.33 (n = 188; 55.3%); C, B.1.91 (n = 9; 2.6%), B.1.195 (n = 3; 0.9%), and B.1.212 (n = 1; 0.3%) (Fig. **1A**). Eight genomes (2.4%) did not cluster within the predicted lineages and were therefore named “others”.

**Fig. 1.**
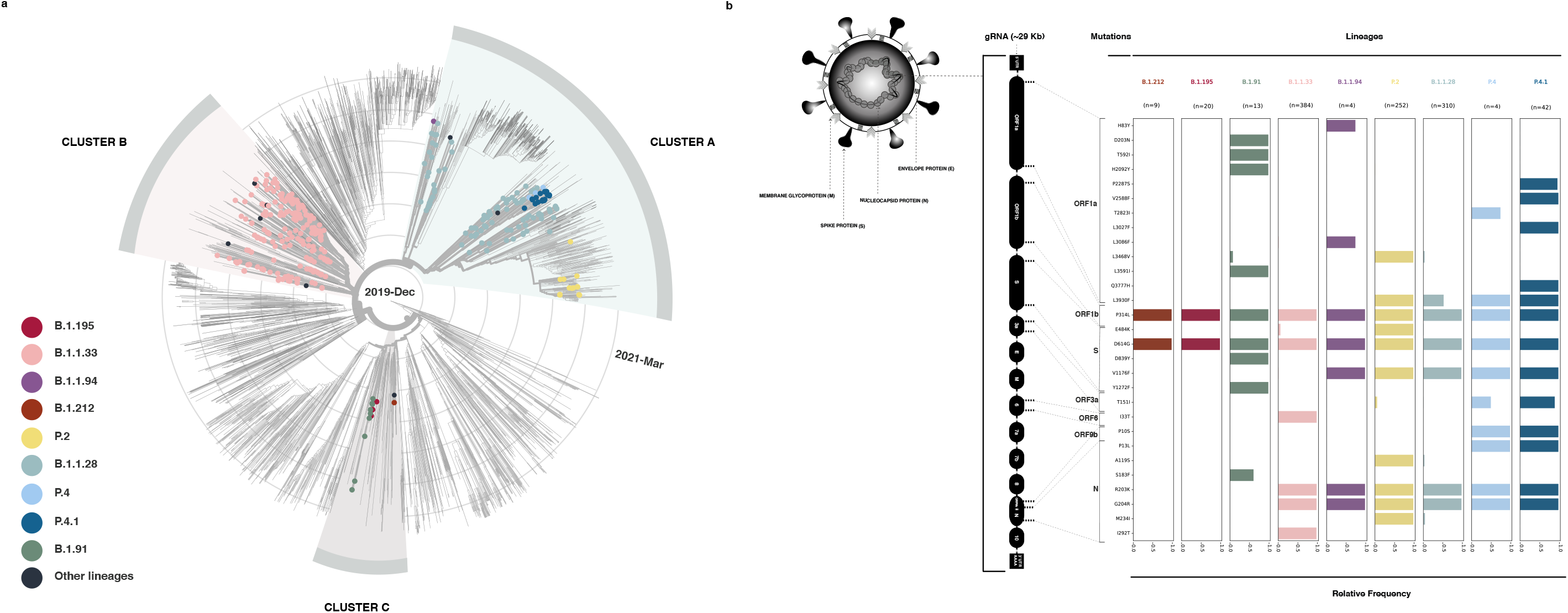
Phylogenomic reconstruction of the SARS-CoV-2 genome. **a** Phylogenetic tree of 340 sequences described in this study combined with a dataset of 3,965 South American genomes. Three main clusters are highlighted: clusters A and B comprise lineages B.1.1.28 and B.1.1.33, respectively, and cluster C is composed of lineages B.1.91, B.1.195 and B.1.212. Branch lengths are proportional to the collection date (between Abril 2020 and March 2021). Concentric circular rings represent the timeline (interval every three months). Each filled circle represents our SARS-CoV-2 sequences, and they are colored according to lineage (legend box). **b** Highlighter plot showing SARS-CoV-2 mutation patterns along the genome evaluated here. Mutations are color-coded according to the lineages shown in Figure 1A legend..

Among all genomes sequenced in this study, we found a total of 503 nonsynonymous mutations compared to the reference Wuhan-Hu-1 sequence. Nonsynonymous mutations were found mainly in ORF1a (211 mutations), ORF1b (91), S (60), ORF3a (39), N (37), ORF8 (24), ORF7a (17), ORF9b (8), M (7), ORF6 (4), E (3), and ORF7b (2) (Supplementary Fig. 3).

Within cluster A, 22 genomes, initially classified as B.1.1.28, clustered together and shared unique amino acid substitutions: ORF3a T151I, ORF9b P10S, and N P13L (Fig. **1B**, Fig. **2**). This putative novel lineage was further investigated in the context of all B.1.1.28 genomes available in GISAID (Supplementary Fig. 4; Supplementary Data 4). Again, a discrete group of genomes presenting the mentioned amino acid changes was identified (Fig. **2**), and it was termed the P.4 lineage. Inside the P.4 clade, we found a distinct cluster, named P.4.1, of sequences presenting four additional unique amino acid changes in ORF1a (P22875S, V2588F, L3027F, Q3777H) and two synonymous mutations in the ORF1a gene (C1288T and G10870T) (Fig. **1B**; Fig. **2**).

**Fig. 2.**
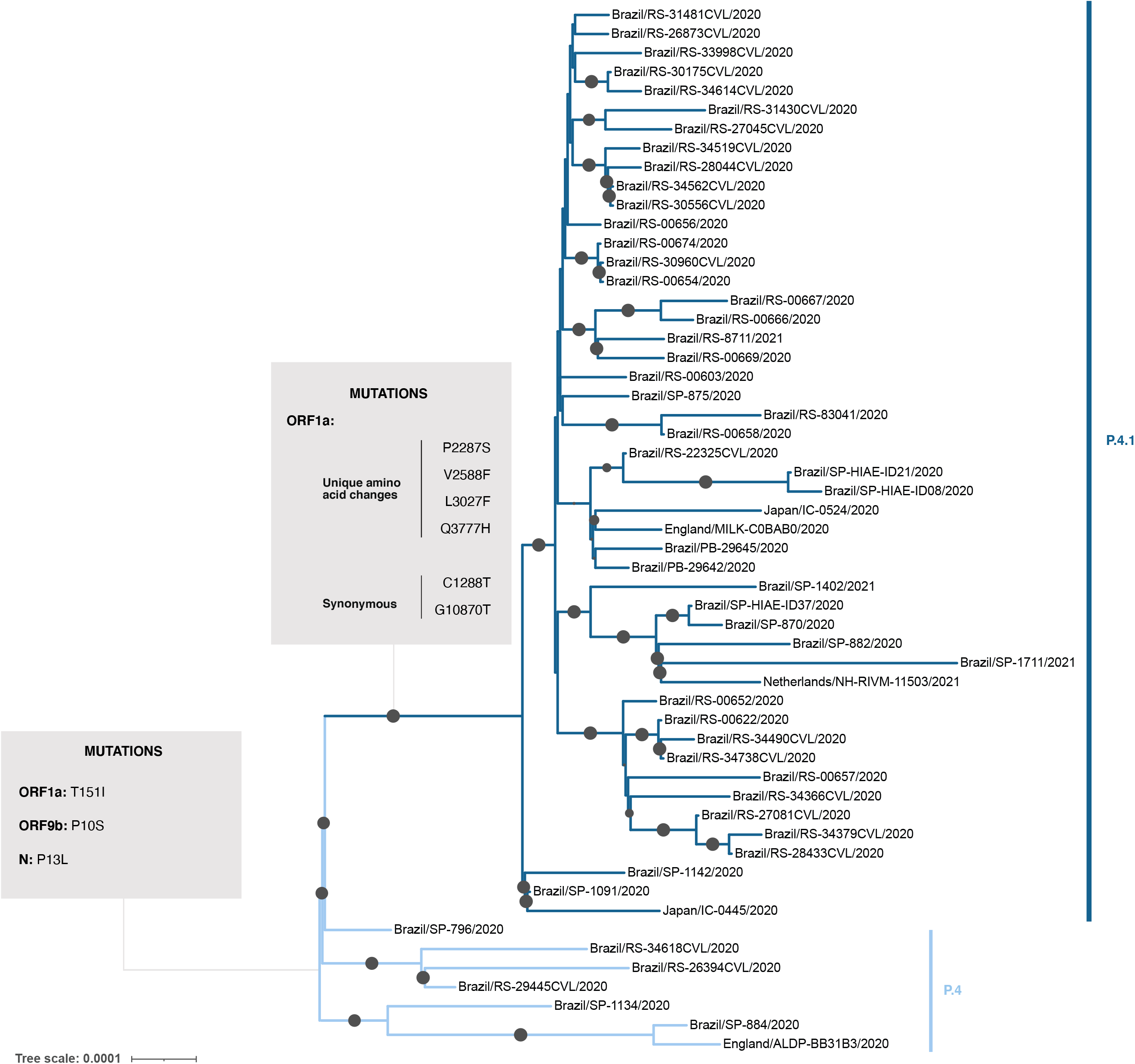
Phylogenomic tree of P.4 and P.4.1 lineages. Phylogenetic inference was performed using all B.1.1.28 sequences available in GISAID. The cluster comprising P.4 and P.4.1 was evident. Mutations shared between P.4 and P.4.1 (in ORFa, ORF9b and N) as well as unique amino acid changes and synonymous mutations to P.4.1 present in ORF1a are highlighted in the boxes. Circles positioned over branches represent ultrafast bootstrap values, and their diameters are proportional to the range from 70 to 100.

Considering the other lineages, it is worth noting that B.1.91 presents two amino acid changes in the spike protein. In turn, all P.2 samples have the E484K, V1176F, and D614G mutations in the spike protein, and all those from Rio Grande do Sul present the synonymous mutation T3766C (Fig. **1B**).

### Temporal and spatial dynamics of SARS-CoV-2 lineages

The nine assigned lineages were investigated regarding their introduction in RS (Supplementary Fig. 5; Supplementary Data 2). Clade A (B.1.1.28, P.2, P.4 and P.4.1 lineages) and clade B (B.1.1.33 lineage) showed multiple introductions from different Brazilian regions (Fig. **1A**; Supplementary Fig. 5A and 5B). On the other hand, clade C comprises the European lineages, B.1.91, B.1.195 and B.1.212. The B.1.91 group is immersed within a European clade. B.1.212 forms a Brazilian group that lies within a clade in which the last common ancestor came from Africa (confidence 70%) (Supplementary Fig. 5C and Supplementary Data 2). In turn, the B.1.195 group is composed of samples obtained in different countries, mostly from South America (Fig. **1A**; Supplementary Fig. 5C; Supplementary Data 2).

The profile of SARS-CoV-2 epidemiological progression over time highlighting main events in the context of Brazil and RS epidemics is shown in Figure 3. At the beginning of the sampled period (April), B.1.1.28 and B.1.1.33 were the predominant lineages circulating in our samples as well as in other Brazilian regions. P.4.1 was first detected in Taquara City on 07 October 2020 (epidemiological week 41), and since then, our findings have shown an obvious rise in the lineage. Based on our phylogenomic analyses, it was possible to infer that P.4.1 emerged between 20 June 2020 and 23 July 2020. The P.4.1 lineage was first detected in a patient who was exposed in Goiás (Brazil/SP-HIAE-ID37/2020), followed by a sequence obtained in São Paulo (Brazil/SP-1091/2020). The emergence and spread of the P.4.1 lineage in South Brazil occurred around the 40th epidemiological week (October 2020). Subsequently, P.2 lineages, already circulating in other regions, showed massive transmission in the community starting on 22 October 2020 (epidemiological week 43) in Porto Alegre, the capital city of RS.

**Fig. 3.**
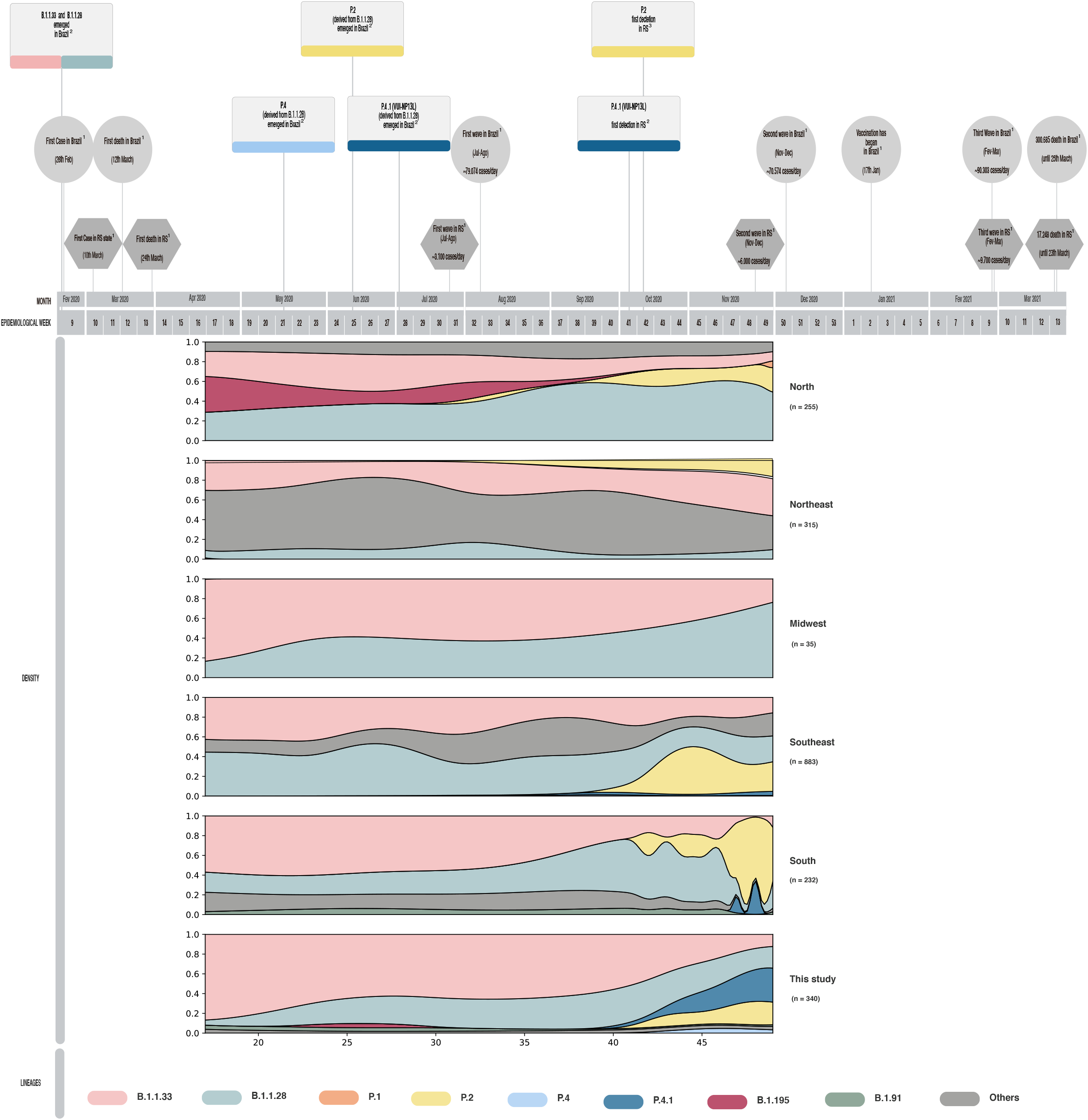
Timeline and density plot of Brazilian SARS-CoV-2 genomes during epidemiologic weeks. The timeline of the main events and circulating lineages of the SARS-CoV-2 pandemic in RS and Brazil from epidemiological weeks 9 of 2020 to 13 of 2021 are highlighted, representing the first year of the pandemic. The frequency of SARS-CoV-2 lineages found in this study in comparison to lineage distribution in other Brazilian regions is shown during our sampling period. **1** Data available in https://covid.saude.gov.br/; **2** Origin dates inferred in this study; **3** GISAID.

The geographic distribution of lineages was evaluated for all RS-sampled regions since the first detection of P.4.1 (Fig. **4**). B.1.1.28, B.1.1.33 and P.4.1 were found across all regions, while the B.1.91 lineage was found only in regions 3 and 2, with a frequency of approximately 4%. We detected lineages P.2 and P.4.1 distributed in 8 and 11 cities, respectively. P.4.1 was the most prevalent lineage circulating in region 2 (36%) and represented 15.4 and 20% of genomes in regions 3 and 1, respectively. On the other hand, P.2 was more frequent than P.4.1 in region 4, but equally frequent in regions 3 and 1.

**Fig. 4.**
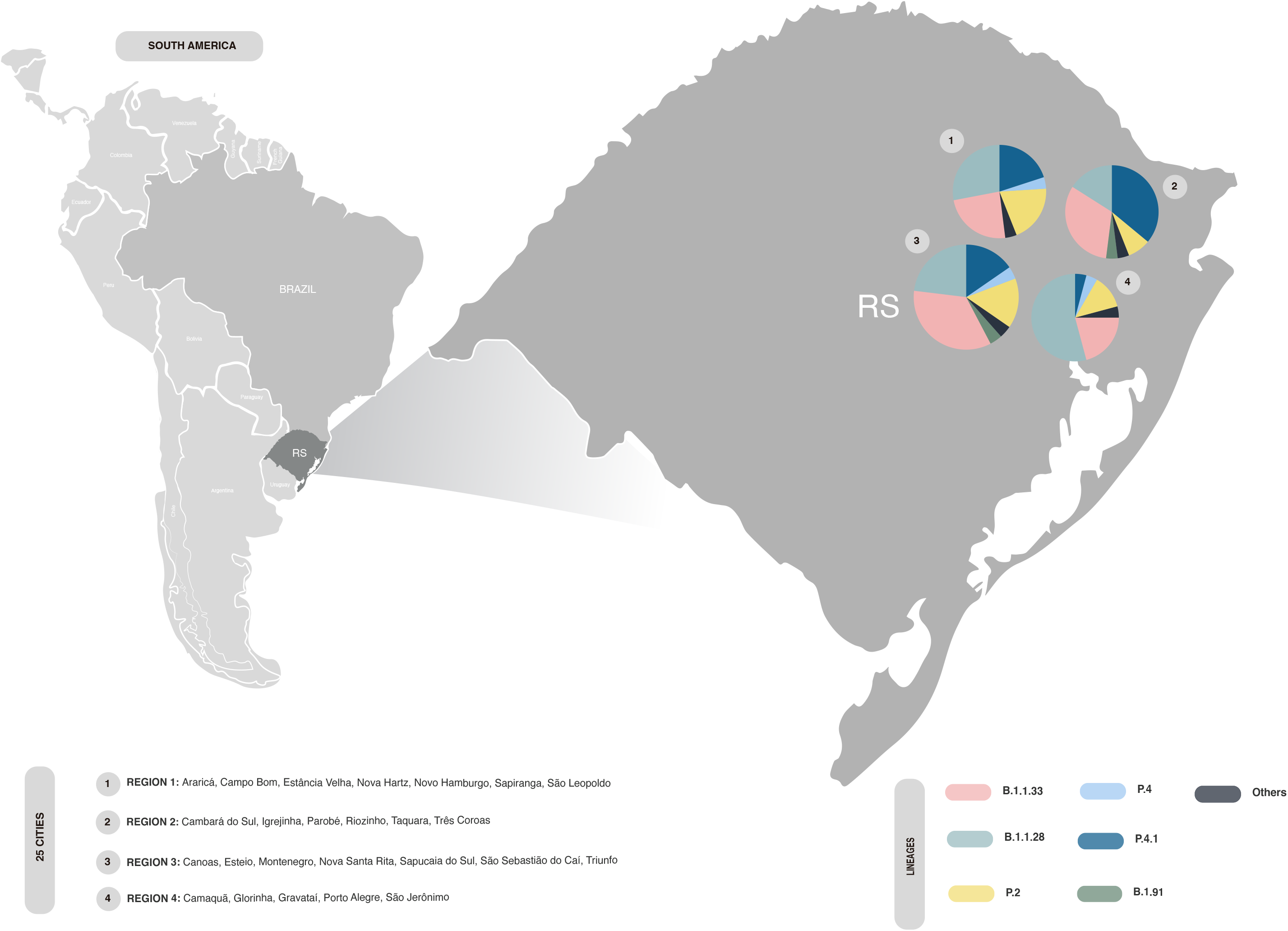
Spatial distribution of the SARS-CoV-2 lineages between 07 October and 30 November 2020 (epidemiological weeks 41 - 49) in four regions of RS. The pie charts represent lineage spreads that are proportional to the number of sequences belonging to each sampled region composed of 25 cities.

## DISCUSSION

This is the largest genomic study of SARS-CoV-2 in Brazil considering the number of sequences and the temporal monitoring window. We characterized two new lineages, P.4 and P.4.1, and identified the massive spread of P.2 and P.4.1 lineages since October 2020 in South Brazil.

The main strength of our study was the analysis of a substantial number of genomes, which represent a geographic region of 33 cities over 8 months. This approach was decisive to evaluate the lineage prevalence across sampled regions and monitor the emergence of new variants of interest. We optimized the sequencing library preparation protocol, improving SARS-CoV-2 genome amplification and enhancing sequencing performance. Although we investigated different regions of the RS, including the metropolitan region, the sampling was done in only one state, which did not allow extrapolation of the findings to the rest of Brazil. Sequences evaluated in our study are restricted to 2020; therefore, we cannot rule out if the lineages are still active across southern Brazil.

Of the seven main circulating lineages, B.1.1.33 and B.1.1.28 were the most predominant, mainly in the initial period of the study. Our findings are supported by other Brazilian sequencing studies, which showed that B.1.1.33 and B.1.1.28 are the most prevalent lineages of the country, including South Brazil^16^.

Regarding phylogeographic analyses, B.1.1.33 and B.1.1.28 lineages were imported from other Brazilian states, and subsequently transmitted among the local community. According to Franceschi et al (2021), B.1.1.33 may have arisen in Rio de Janeiro and disseminated across Brazil^16,18,20,23^. This lineage was also associated with secondary outbreaks in Argentina and Uruguay, countries bordering RS^18,24^.

Our phylogenomic analyses provided evidence that B.1.1.28 is a paraphyletic group composed of several subclades of distinct evolutionary origins and unique genetic signatures. Therefore, we found that the B.1.1.28 lineage may have arrived in RS during multiple episodes, although Francisco Jr. et al (2021) suggested that it was introduced by a single seeding event^17^. Despite most lineages having been introduced from other Brazilian states, it seems that B.1.91 was directly imported from Europe. However, it is worth noting that for reliable geographic transmission inference, careful sampling is necessary. Available genome samples in GISAID are unbalanced among different Brazilian regions, which may lead to incorrect inferences about virus origins.

Our results also indicated a sharp increase in the P.2 lineage almost concurrently with P.4.1. Recently, Lamarca et al (2021) reported P.2 as the most abundant lineage in Northeast and Southeast Brazil. Furthermore, the authors revealed the beginning of its circulation in March 2020, exhibiting an intense transmission between December 2020 and January 2021^23^. Precise detection of the origins of an outbreak succeeds only when the genetic background of the pathogen is known. Between 16 August 2020 and 14 November 20 (epidemiological weeks 34-46), only 19 SARS-CoV-2 sequences from South Brazil were available in GISAID. In this sense, our dataset was crucial for tracing the beginnings of an outbreak in South Brazil caused by a novel SARS-CoV-2 lineage, which here was termed P.4.1.

Francisco Jr. et al. (2021) independently detected P.4.1, provisionally naming the lineage as VUI-NP13L^17^. They also pointed out that this lineage is characterized by the P13L amino acid change in the N protein. However, in this study we demonstrated that at least two viral sublineages possess this modification, and we described their genetic signatures. These lineages were named according to the criteria described by Rambault et al. (2021), in which names for descendants of the B.1.1.28 lineage cannot exceed three sublevels in the current classification system, thus giving P.4 and P.4.1^21,25^.

Although P.4.1 probably emerged in Goiás or São Paulo around Jun-Jul 2020, this lineage was only identified in South Brazil at the beginning of October 2020. According to an independent phylodynamic reconstruction, P.4.1 rapidly arrived in the southeastern and northeastern regions of Brazil and seems to have been exported to Japan, the Netherlands and England^23^. The success of the spread of P.4.1, beyond having important mutations in the spike protein (V1176F, D614G), also shared by P.2 and P.1 lineages, could be justified by the unique amino acid changes in ORF1a^16,26,27^. This gene is known to encode a polyprotein involved in the replication complex^28^. Previous studies concerning SARS-CoV and MERS indicated the role of ORF1a in survival and adaptation to the host^28,29^. Further, Forni et al. (2016) suggested that ORF1a positive selection might contribute to host shifts or immune evasion^29^. However, we cannot discard the increase in frequency of P.4.1 due to random causes such as a “founder effect,” regardless of the mutation effects of ORF1a on SARS-CoV-2 fitness^27^.

Our study reinforces the importance of consistent and continuous genomic surveillance for evaluating the genomic background of SARS-CoV-2 in a given spatiotemporal setup. These data are fundamental for inferring SARS-CoV-2 outbreaks and revealing signatures, activity and origins of the lineages. Furthermore, genome surveillance is an invaluable resource to guide decision making of the Brazilian Public Healthcare System. Immunization campaigns could be especially affected by the emergence of novel lineages that evade antibodies generated by current vaccines. Future studies are needed to assess the fate of P.2 and P.4.1 over time, and if they are still observed, we need to evaluate the impact of amino acid changes on the fitness of these lineages.

## METHODS

### 1. Bioethics, sample collection and processing

The study was approved by the Institutional Review Board of Moinhos de Vento Hospital under protocol number 32149620.9.0000.5330. The Clinical Epidemiology Laboratory (Epiclin), located in the Federal University of Health Sciences of Porto Alegre (UFCSPA) and supported by Moinhos de Vento Hospital, was responsible for conducting the diagnostic tests of SARS-CoV-2 in four regions belonging to the Northeast and the Metropolitan Mesoregions of the state of Rio Grande do Sul, southern Brazil. The samples sequenced in this study were collected in these four regions, covering a total of 33 municipalities described in Supplementary Data 1.

Diagnostic testing for SARS-CoV-2 using RT-PCR started in April 2020 and was completed in December of the same year. The total number of clinical samples received by the Epiclin Laboratory was 33,788 resulting in 31,318 individuals tested. Of the total tests carried out, 13,701 were positive for SARS-CoV-2 (40.55%).

Nasopharyngeal and oropharyngeal samples from SARS-CoV-2-infected individuals were collected according to the CDC guidelines^30^. Total nucleic acid was extracted from the samples using the MagNA Pure LC Total Nucleic Acid Kit -High Performance in the MagNA Pure Instrument (Roche) or MagMaxTM Viral/Pathogen (MVP III) Nucleic Acid Isolation kit (Applied Biosystems) in the KingFisher Flex System (Thermo Fisher Scientific). These samples were evaluated by real-time RT-PCR multiplex Allplex SARS-CoV-2 (Seegene). Nucleic acid samples were immediately stored at -80 °C.

### 2. Study design

Total nucleic acids from positive samples of the Epiclin Laboratory diagnostic tests were selected considering a spatiotemporal outlook. Strata were defined using two criteria: epidemiological week and residence of the participant. The selected period includes epidemiological weeks between 17 (April 2020) and 49 (November 2020) from four regions (Novo Hamburgo, Taquara, Canoas, and Porto Alegre). In each of the 132 strata (33 weeks and 4 regions), samples were selected randomly for sequencing, totaling 353 RNA samples (Supplementary Methods).

### 3. Library preparation and sequencing

Libraries were prepared using the QIASEQ SARS-CoV-2 Primer Panel (Qiagen) and QIAseq FX DNA Library CDI Kit (Qiagen) according to the manufacturer’s instructions, except for the annealing temperature of the primers. The QIASEQ SARS-CoV-2 Primer Panel contains a PCR primer set for whole-genome amplification of SARS-CoV-2 whose primer sequences were based on the ARTIC network nCoV-2019. To improve amplification of amplicon 64, the annealing temperature of primers was reduced by 1 (one) degree Celsius, from 65 °C to 64 °C (Supplementary Fig. 1). The libraries were quantified using the QubitTM dsDNA HS Assay kit on a Qubit 4.0 fluorometer (Thermo Fisher Scientific) and normalized to equimolar concentrations. A pool of all of the normalized libraries was prepared and diluted to a final concentration of 10 pM and sequenced on the Illumina MiSeq platform using the MiSeq Reagent Kit v3 600 cycles (Illumina).

### 4. Genome assembly

Raw paired-end reads were processed using a bioinformatic pipeline previously described with some modifications^31, 32^. The reads were mapped with BWA 0.7.17 software^33^ to the Wuhan-Hu-1 reference genome (NC_045512.2) and converted to BAM format using samtools v1.7^34^. The primer sequences were trimmed with the iVar v.1.2.3 package, and consensus sequences were generated considering a Phred quality score minimum of 20 and N for regions with coverage depths less than 10 bases. The quality of the genome sequences was assessed using Nextclade version v. 0.14.1. Sequences classified as “bad” in the overall quality evaluation were discarded from further analyses. Viral genomes were deposited in GISAID, and the accession numbers are available in Supplementary Data 1^33^.

### 5. Identification of SARS-CoV-2 lineages and mutations

Viral lineages were identified with PANGOLIN (Phylogenetic Assignment of Named Global Outbreak LINeages) version 2.3 (pangoLEARN version 2021-02-12). Nucleotide and amino acid mutations were mapped using Nextclade version v 0.14.1.

Novel lineages were identified following the approach described in Rambaut et al., 2020. All genome sequences of the Pangolin lineages B.1.1.28 were recovered from the nextfasta file available in the GISAID database (build 2021-03-08) (Supplementary Data 4). Genome sequences obtained in this study were concatenated with global sequences according to the Pangolin lineage. Sequences were filtered using Augur Filter subcommand with a minimal length of 27,000 nucleotides, and subsequently aligned with MAFFT, FFT-NS-2 option with default parameters. The phylogenetic tree was built using IQ-Tree using the GTR model and ultrafast bootstrapping with 1,000 replicates, and subsequently visualized in iTOL v6^35^.

### 6. Phylogenomic, phylogeographic, and phylodynamics analyses

Phylogeographic and phylodynamic analyses were carried out using Nextstrain, a suite of tools that includes subsampling, alignment, phylogenetic reconstruction, geographic and ancestral trait reconstruction, and inference of transmission events^36^. For these analyses, our dataset of 340 local genome sequences was concatenated with the Nextstrain South America dataset (build 2021-03-03) (Supplementary Data 2 and 3). All 3,965 sequences were included in the pipeline (subsampling step was bypassed). Samples were classified according to their exposure location, taking into consideration the following rationale: samples from Rio Grande do Sul, samples from other regions of Brazil, samples from South America, excluding Brazilian sequences; and samples from other continents. Trait reconstruction was performed considering this custom geographic trait. Kernel density of the lineages during the epidemiological weeks was built using a script written in Python (Seaborn library).

## Data Availability

The data that support the findings of this study are openly available in GISAID repository.

## Data availability

The 340 SARS-CoV-2 sequences obtained in this study were submitted to the GISAID portal and are available in Supplementary Data 1.

## Acknowledgments

We thank the Federal University of Health Sciences of Porto Alegre (UFCSPA), which provided post doctoral financial support to APMV. We also thank Dr. Vanessa Mattevi, Dr. Graziela Agnes, Dr. Carmela Tagliari, Dr. Marília Zandoná, Dr. Augusto Bacelo Bidinotto, Lara Garcia, Tiago Fetzner, and Milena Mantelli for providing technical support. We thank all the authors who have shared genome data on GISAID, and we have included a table (Supplementary Data 5) listing the authors and institutes involved.

## Author contributions

FHS helped with study design, sequencing, performed the bioinformatic analysis, data interpretation, and writing; APMV and JP helped with study design, performed the sequencing, data interpretation, figure drawing, and writing; JC and CBC helped with study design, data interpretation and writing; VSR helped with sampling design; AS, FM and GPA revised the manuscript; EMW conceived the study and revised the manuscript. All authors contributed to the article and approved the submitted version.

