## Supplementary Fig. for "Emergence of the novel SARS-CoV-2 lineage P.4.1 and massive spread of P.2 in South Brazil"

Supplementary Data 1. **Metadata of the 340 whole-genome sequences obtained in this study.**

Supplementary Data 2. **Interactive phylodynamic and phylogenomic data of SARS-CoV-2.** Sequences of this study were concatenated with the South America dataset. To visualize the Nextstrain build, the JSON file was loaded at the following link: https://auspice.us/. The sequences of our study were highlighted by filtering according to the following authors: "Sant'Anna et al". To visualize the phylodynamics in Rio Grande do Sul (Supplementary Figure 4), the tree was colored by "custom_location".

Supplementary Data 3. **South America dataset of SARS-CoV-2 genomes (n = 3,965) retrieved from GISAID enriched for South America and used as a reference for the construction of the phylogeny, as well as their respective accession numbers and corresponding lineages.**

Supplementary Data 4. **Dataset of B.1.1.28 sequences from GISAID used for phylogenomic analysis.**

Supplementary Data 5. **Acknowledgement table.**

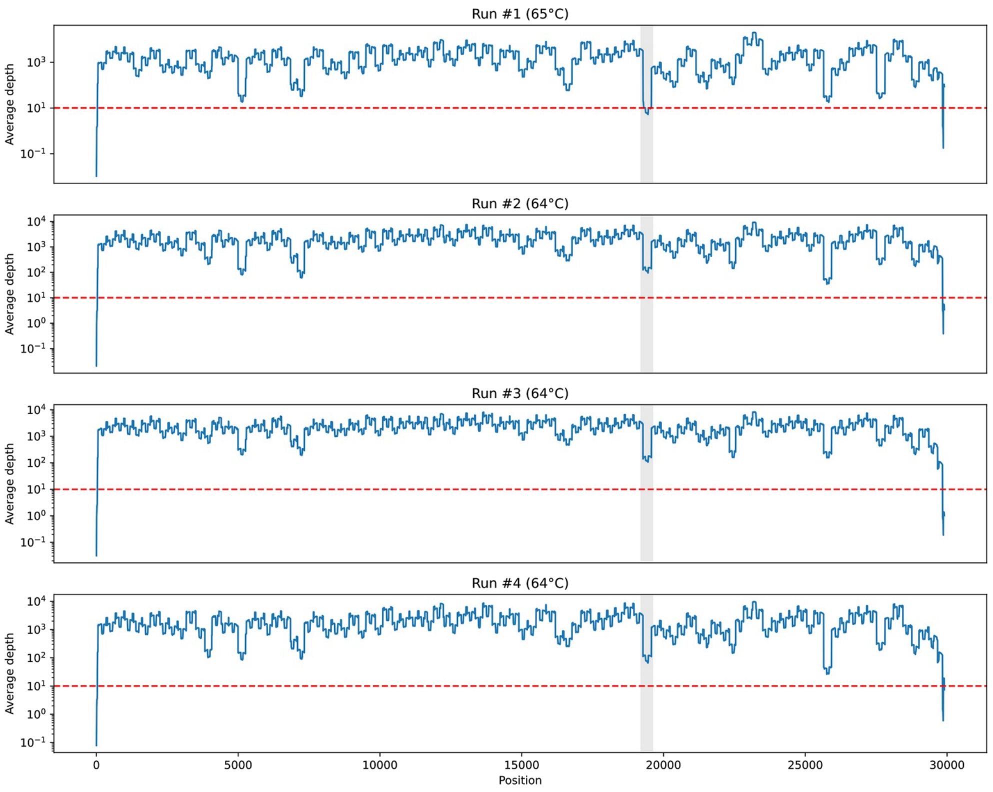


Supplementary Fig. 1. **SARS-CoV-2 genome amplification performance by target position using two different annealing temperatures.** Plots of the depth coverage of the SARS-CoV-2 genomes per position in the different sequencing runs. The first plot represents the mapped genome profile with annealing temperature at 65 °C in the multiplex tiling-PCR, and the other three show the profile at 64 °C. The gray stripe highlights the region spanning the position 19204 to 19616, corresponding to amplicon 64 of the ARTIC framework.

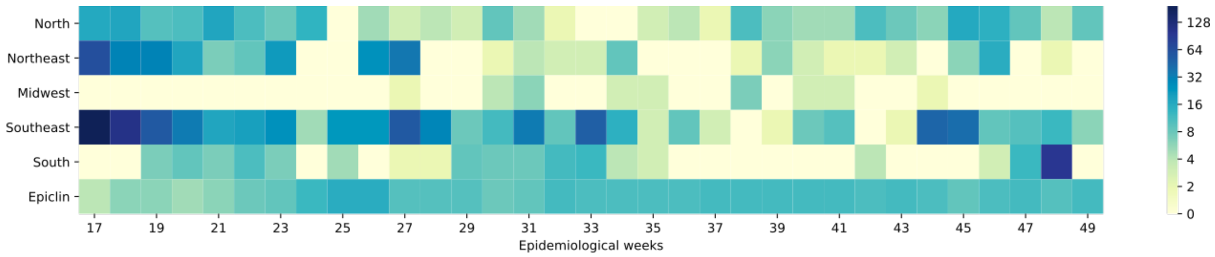


Supplementary Fig. 2. **Distribution of SARS-CoV-2 genomes in Brazilian regions and Epiclin from April to November (epidemiological weeks 17 to 49).** Heatmap showing SARS-CoV-2 genomes in the five Brazilian regions compared to the samples sequenced in this study between epidemiological weeks 17 and 49. Brazilian regions are in the rows and epidemiological weeks are in the columns. The relative abundance is represented by colors (white, lowest abundance; dark blue, highest abundance), as indicated in the legend. Average number and the standard deviation of genomes per week: North, 7.72 ± 5.26; Northeast, 9.54 ± 14.37; Midwest, 1.06 ± 1.08; Southeast, 26.75 ± 37.42; South, 7.03 ± 16.68; Epiclin, 10.30 ± 2.7.

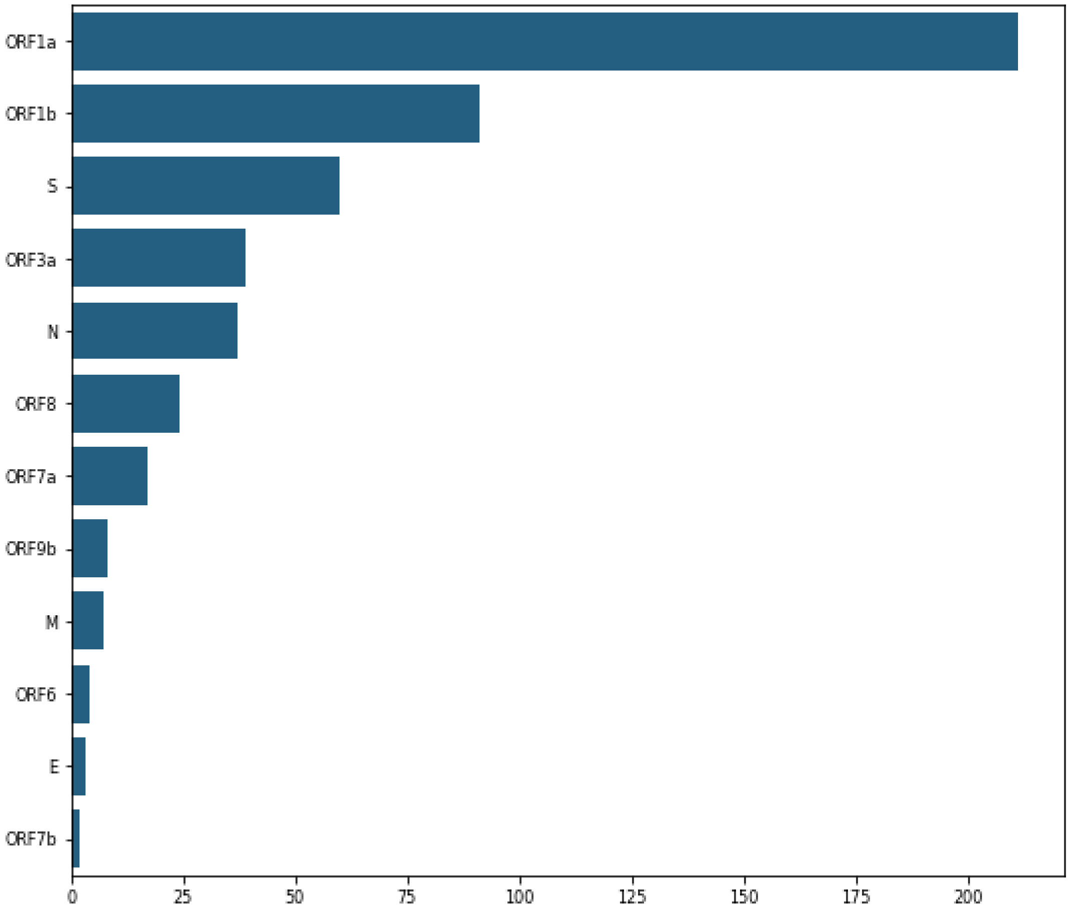


Supplementary Fig. 3. **Frequency of amino acid changes of SARS-CoV-2 proteins.**

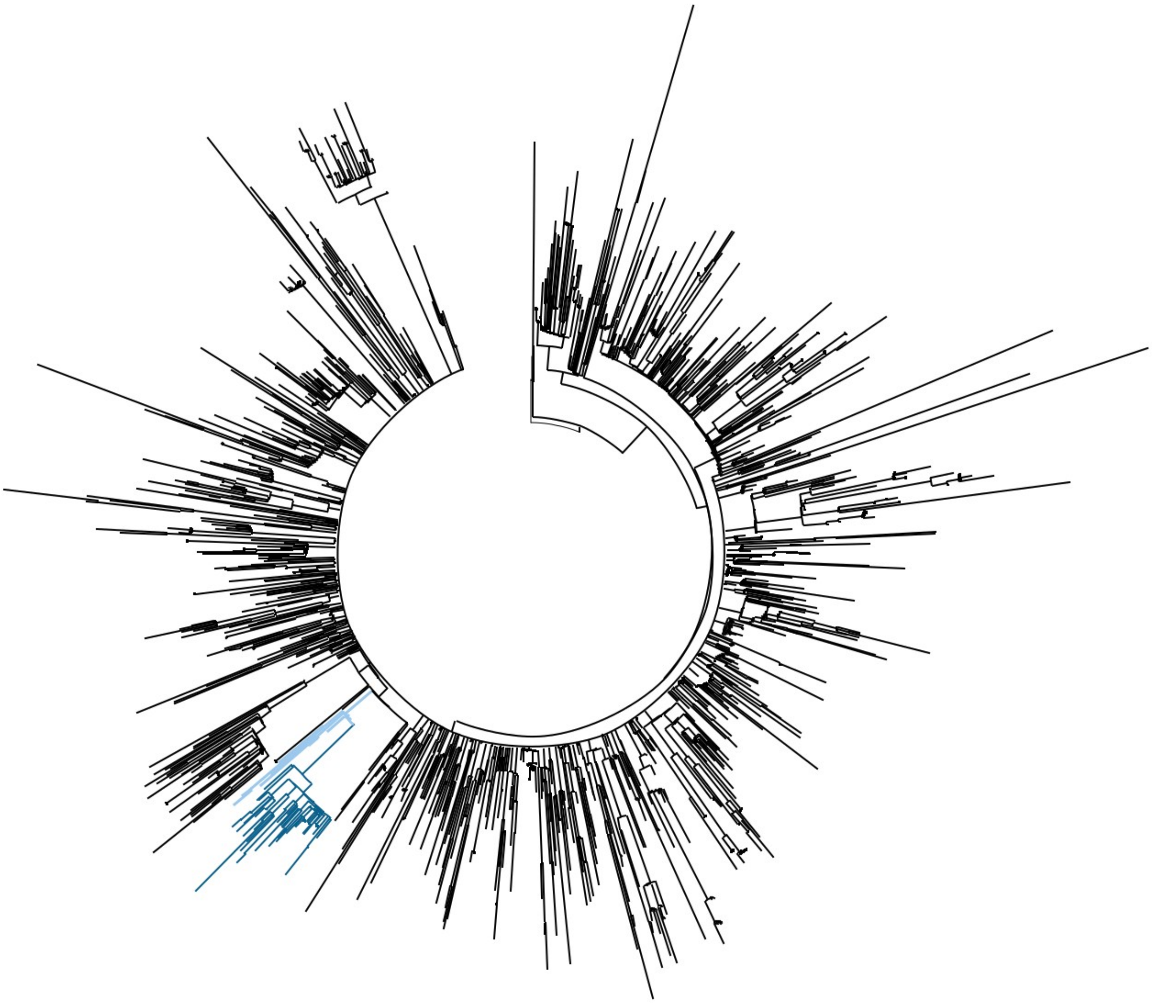

Supplementary Fig. 4. **Phylogenomic reconstruction of the SARS-CoV-2 B.1.1.28 lineage available in GISAID.** Clusters of novel lineages, P.4 and P.4.1, are shown in light and dark blue colors, and other sequences of the B.1.1.28 lineage are shown in black. The branch of novel lineages is supported by a bootstrap of 100%. The phylogeny was estimated with 1,000 bootstrap replicates.

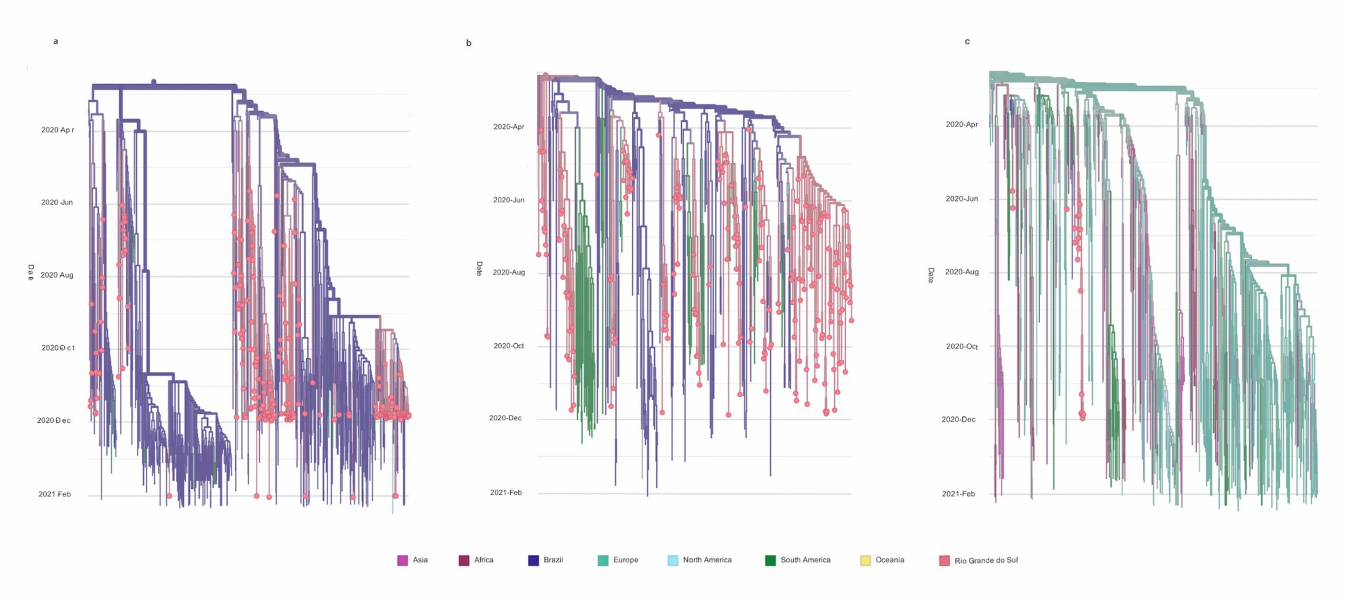


Supplementary Fig. 5. **Introductions of SARS-CoV-2 in Rio Grande do Sul. a** Cluster A from Fig. 1A; **b** Cluster B from Fig. 1A; **c** Cluster C from Fig. 1A. Clades are colored according to the location of their last common ancestor (legend box). Circles depict samples from Rio Grande do Sul.

### Supplementary Methods

#### Sampling criteria

The samples sequenced in this study are from four main regions named regions 1 to 4: Novo Hamburgo (region 1), Taquara (region 2), Canoas (region 3), and Porto Alegre (region 4). These regions are located in the State of Rio Grande do Sul in southern Brazil. Each region comprises following cities: Novo Hamburgo covered 9 cities (Araricá, Campo Bom, Dois Irmãos, Estância Velha, Nova Hartz, Novo Hamburgo, Portão, São Leopoldo and Sapiranga); Taquara included 7 cities (Cambará do Sul, Igrejinha, Parobé, Riozinho, Rolante, Taquara and Três Coroas); Canoas included 10 cities (Canoas, Esteio, Harmonia, Montenegro, Nova Santa Rita, Pareci Novo, São Sebastião do Caí, Sapucaia do Sul and Triunfo, Vera Cruz); and Porto Alegre included 7 cities (Porto Alegre, Gravataí, Cachoeirinha, Camaquã, Alvorada, São Jerônimo, Glorinha), total 33 cities that represent the northeast and the metropolitan region of the State.
